# Safe Non-opioid and Non-barbiturate containing Acute Medication Combinations in Headache: An Exhaustive Approach Based on DrugBank Database

**DOI:** 10.1101/2020.06.26.20140897

**Authors:** Victor Kaytser, Pengfei Zhang

## Abstract

**Background:** Rational polypharmacy in abortive medications use is often inevitable for patients with refractory headaches.

**Objective:** We seek to enumerate an exhaustive list of headaches abortive medications that are without drug-drug interactions.

**Methods:** We updated a list of acute medications based on the widely used Jefferson Headache Manual with novel abortive medications including ubrogepant, lasmiditan, and rimegepant. Opioids and barbiturate containing products are excluded. We then conducted an exhaustive search of all pair-wise interactions for this list of medication via DrugBank API. Using this interaction list, we filtered all possible two, three, and four drug combinations of abortive medications. The resultant list of medication was then reapplied to DrugBank to verify the lack of known drug-drug interaction.

**Results:** There are 192 medication combinations that do not contain any drug-drug interactions. Most common elements in these combinations are ubrogepant, prochlorperazine, followed by tizanidine. There are 67 three-drug combinations that do not contain interaction. Only 2 of the four-drug combinations do not yield some form of drug-drug interactions.

**Conclusion:** This list of headaches abortive medications without drug-drug interactions is a useful tool for clinicians seeking to more effectively manage refractory headaches.

## Introduction

Non-opioid and non-barbiturate containing acute therapies are staples of headache management.^1^ However, excessive use of analgesics can often lead to medication overuse headaches.^2^ Therefore practitioners often utilize rational polypharmacy in order to manage refractory patients in real life clinical settings.^3^

Although numerous articles have been published regarding medication interactions in headache medicine, we are unable to find a study conducting an exhaustive search for all medication combinations that are without interactions. We speculate that this is likely due to the heavy computational demands of achieving such an objective. However, this task is now feasible with the advent of application programming interface (API) for online databases. The goal of this study is precisely to derive all possible combinations of abortive medications that are without interactions using DrugBank, an online pharmaceutical database.^4^ We believe that such a list would provide headache providers with an accessible tool for rational polypharmacy.

## Methods

In order to include a broad list of commonly used abortive medications, we included all medications listed in the acute therapy chapter of Jefferson Headache Manual, an influential headache handbook.^5^ (Specifically, Tables 5.5, 5.7, 5.8, 5.14, 5.16) Since abortive medications are often used concurrently with “bridge” therapies, “bridge” medications are included in our list as well. We excluded Midrin, as it is no longer available for prescription in the United States. Since we are interested in non-opioid, non-barbiturate containing products, we also excluded butalbital and opioids. Lastly, we included lasmiditan, ubrogepant, and rimegepant; these mediations received FDA approval after the publication of the Jefferson Manual. Our full list of medications is presented in Table 1. Using this list, we derived all possible combinations of acute medications, paired two at a time. We then algorithmically accessed DrugBank API to determine which pairing yields a pharmacological interaction. We call the result list *c*.

**Table 1:** List of abortives included and # of inclusions.

|  |  |
| --- | --- |
| ubrogepant | 96 |
| prochlorperazine | 41 |
| tizanidine | 37 |
| prednisone | 31 |
| lasmiditan | 28 |
| olanzapine | 23 |
| methylergonovine | 17 |
| acetaminophen | 15 |
| aspirin | 8 |
| diflunisal | 8 |
| fenoprofen | 8 |
| ibuprofen | 8 |
| ketoprofen | 8 |
| meclofenamate | 8 |
| oxaprozin | 8 |
| salsalate | 8 |
| sulindac | 8 |
| tolmetin | 8 |
| etodolac | 6 |
| flurbiprofen | 6 |
| indomethacin | 6 |
| ketorolac | 6 |
| meloxicam | 6 |
| naproxen | 6 |
| piroxicam | 6 |
| droperidol | 5 |
| naratriptan | 5 |
| diclofenac | 4 |
| nabumetone | 4 |
| celecoxib | 3 |
| dihydroergotamine | 3 |
| eletriptan | 3 |
| frovatriptan | 3 |
| metoclopramide | 3 |
| promethazine | 3 |
| sumatriptan | 3 |
| zolmitriptan | 3 |
| almotriptan | 1 |
| chlorpromazine | 1 |
| rizatriptan | 1 |
| ergotamine | 0 |
| haloperidol | 0 |
| dexamethasone | 0 |
| methylprednisolone | 0 |

Using custom software, we then derived all possible combinations of abortive medications when taken two, three, and four at a time. The number of such combinations can be described mathematically by _*a*_C_*2, a*_C_*3*_, and _*a*_C_*4*_, respectively. Here *a* is the number of abortive medications in our list. We call each of these lists L_2_, L_3_, and L_4_, respectively.

Using list *c*, we filtered out elements of L_2_, L_3_, and L_4_ that contain any pairing of interacting medications. This produces a list of medications that are without any drug-drug interaction with respect to each other. This list is presented in Table 2.

**Table 2.**
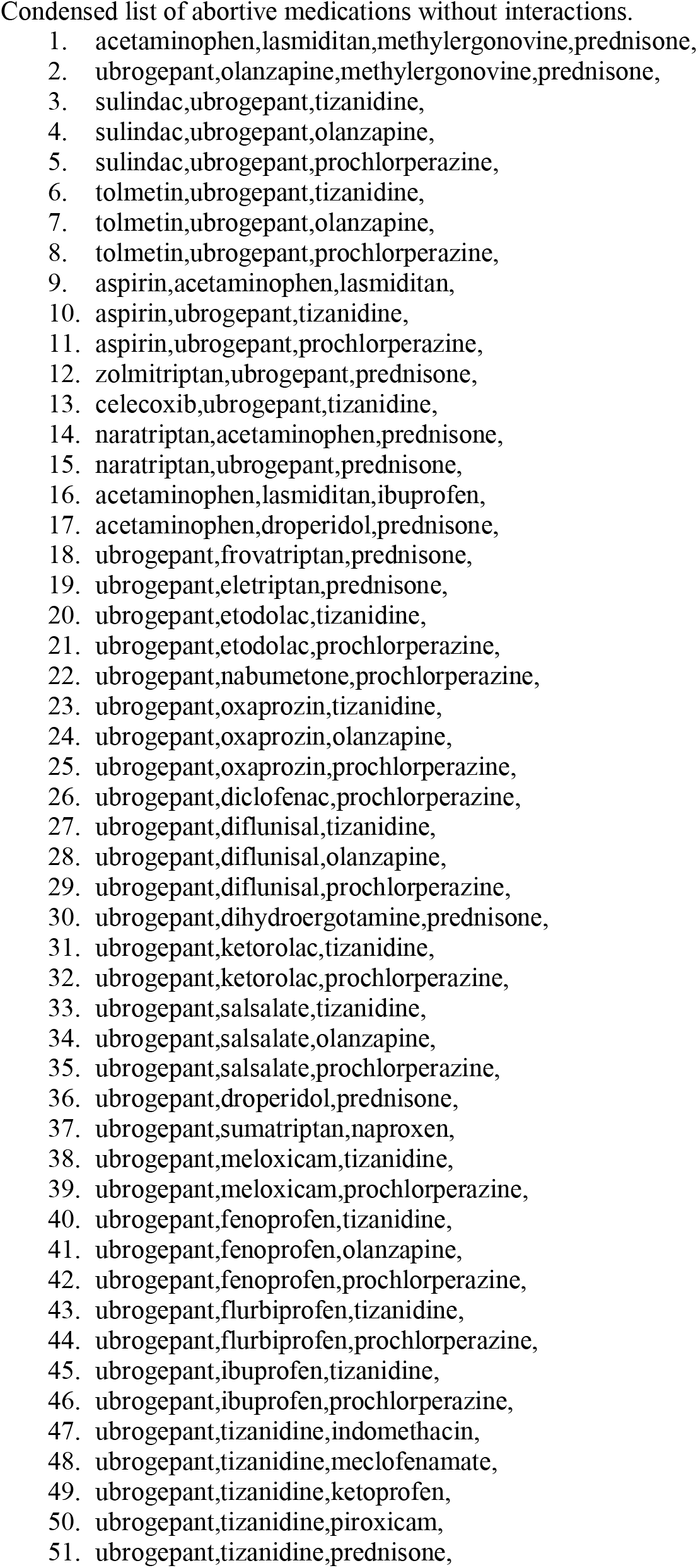
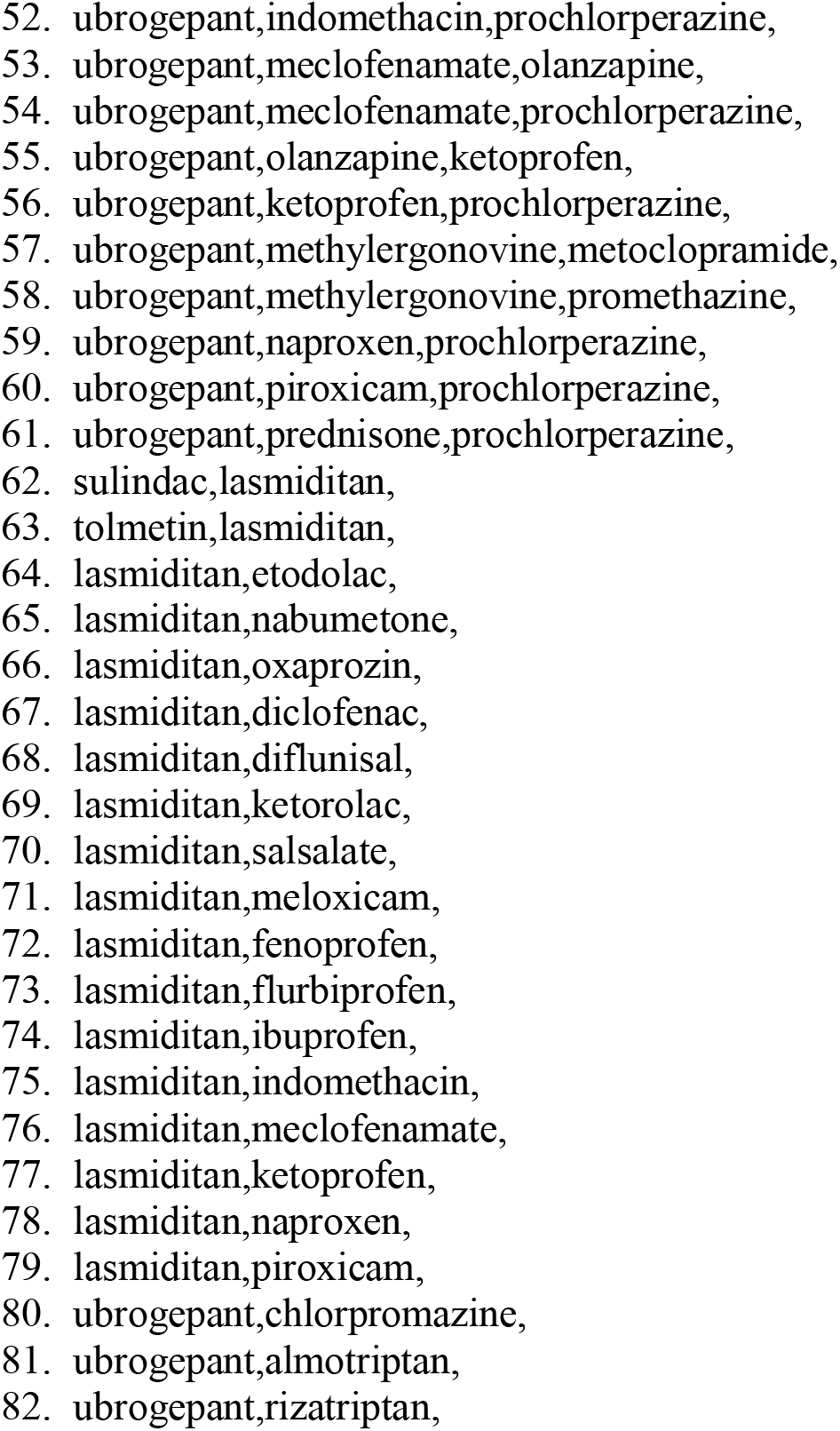

**Table 3:**
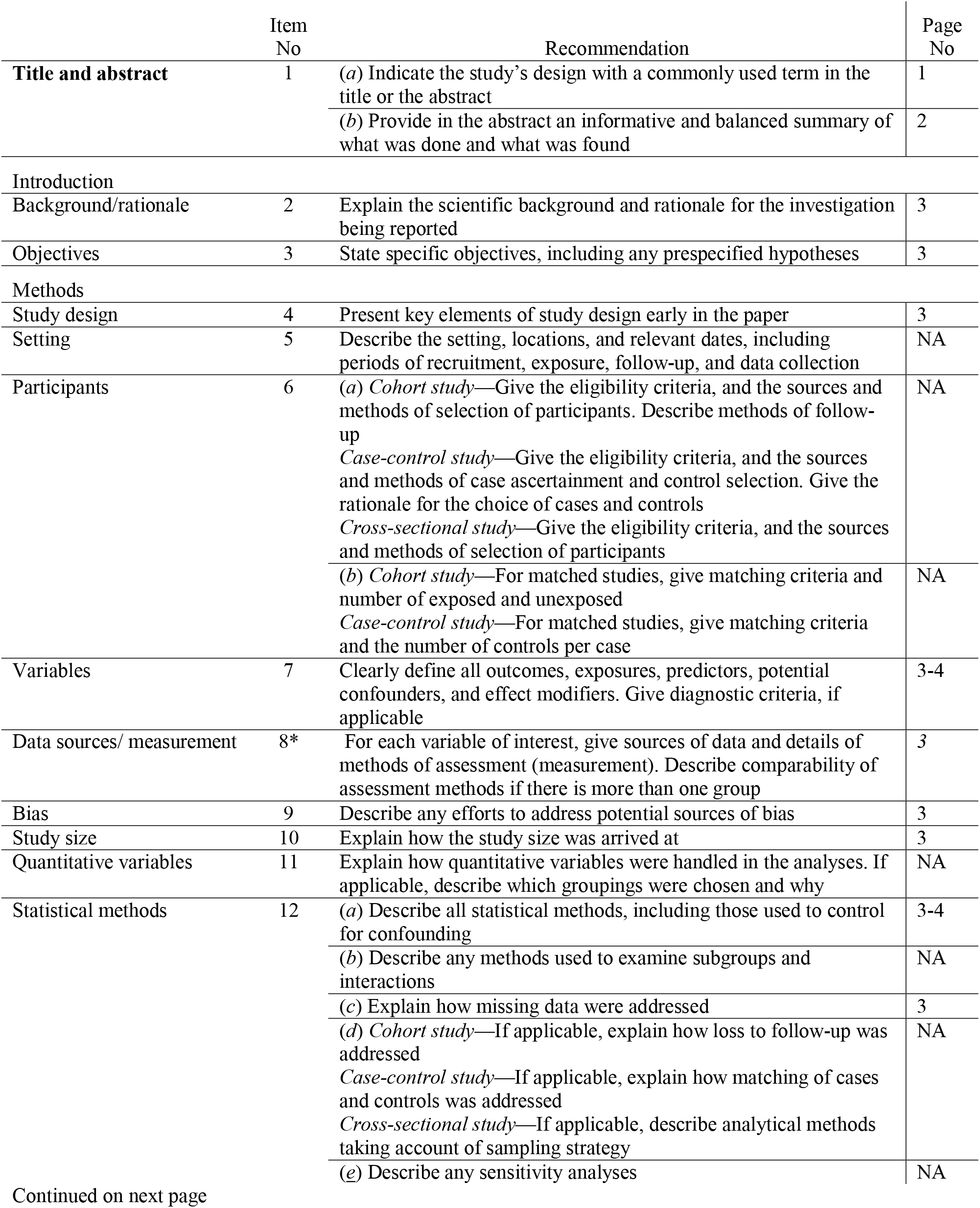
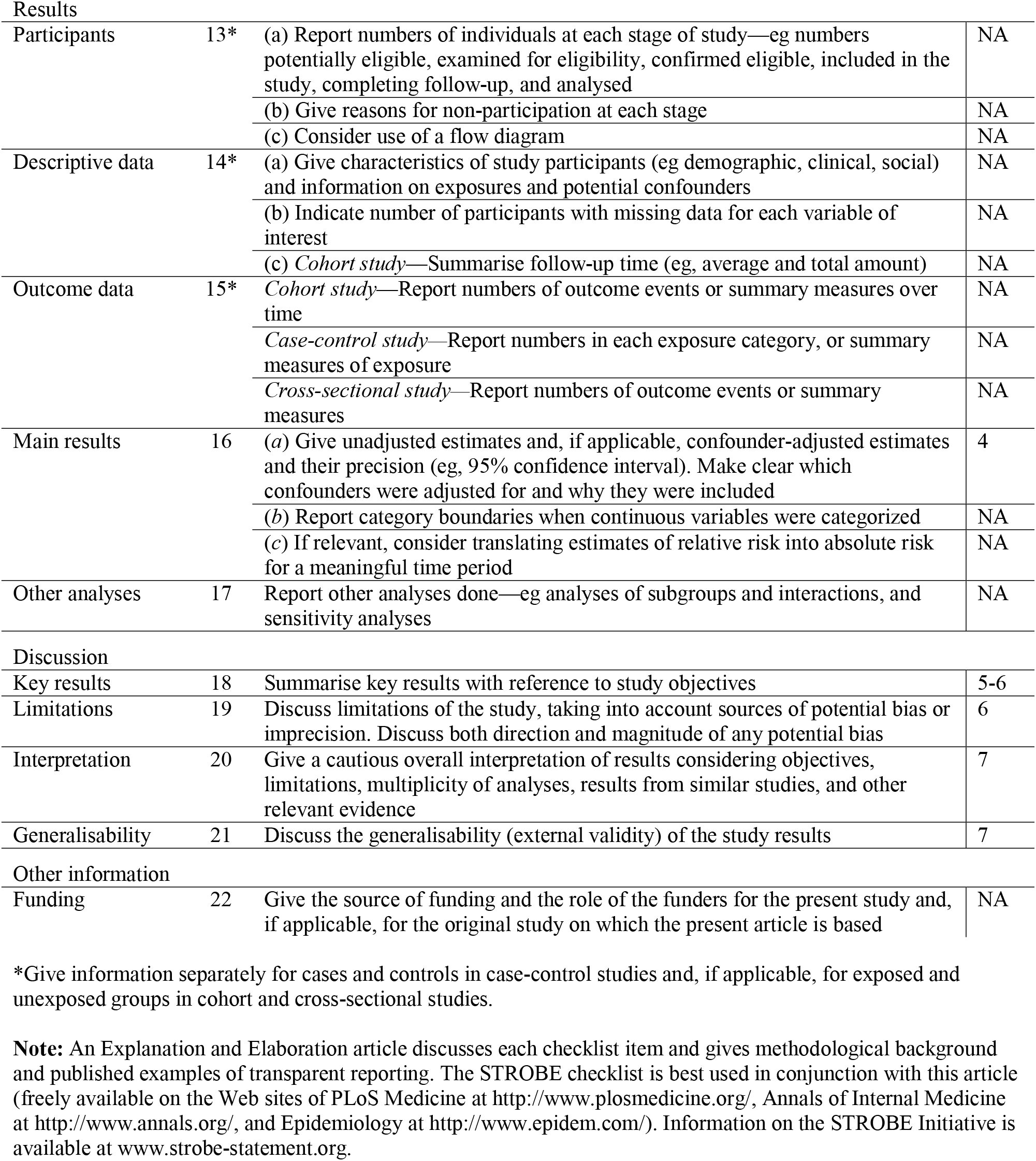
STROBE Statement—checklist of items that should be included in reports of observational studies

|  | Item No | Recommendation | Page No |
| --- | --- | --- | --- |
| Title and abstract | 1 | (a) Indicate the study’s design with a commonly used term in the title or the abstract | 1 |
|  |  | (b) Provide in the abstract an informative and balanced summary of what was done and what was found | 2 |
| Introduction |  |  |  |
| Background/rationale | 2 | Explain the scientific background and rationale for the investigation being reported | 3 |
| Objectives | 3 | State specific objectives, including any prespecified hypotheses | 3 |
| Methods |  |  |  |
| Study design | 4 | Present key elements of study design early in the paper | 3 |
| Setting | 5 | Describe the setting, locations, and relevant dates, including periods of recruitment, exposure, follow-up, and data collection | NA |
| Participants | 6 | (a) Cohort study—Give the eligibility criteria, and the sources and methods of selection of participants. Describe methods of follow-up | NA |
|  |  | Case-control study—Give the eligibility criteria, and the sources and methods of case ascertainment and control selection. Give the rationale for the choice of cases and controls |  |
|  |  | Cross-sectional study—Give the eligibility criteria, and the sources and methods of selection of participants |  |
|  |  | (b) Cohort study—For matched studies, give matching criteria and number of exposed and unexposed | NA |
|  |  | Case-control study—For matched studies, give matching criteria and the number of controls per case |  |
| Variables | 7 | Clearly define all outcomes, exposures, predictors, potential confounders, and effect modifiers. Give diagnostic criteria, if applicable | 3-4 |
| Data sources/ measurement | 8* | For each variable of interest, give sources of data and details of methods of assessment (measurement). Describe comparability of assessment methods if there is more than one group | 3 |
| Bias | 9 | Describe any efforts to address potential sources of bias | 3 |
| Study size | 10 | Explain how the study size was arrived at | 3 |
| Quantitative variables | 11 | Explain how quantitative variables were handled in the analyses. If applicable, describe which groupings were chosen and why | NA |
| Statistical methods | 12 | (a) Describe all statistical methods, including those used to control for confounding | 3-4 |
|  |  | (b) Describe any methods used to examine subgroups and interactions | NA |
|  |  | (c) Explain how missing data were addressed | 3 |
|  |  | (d) Cohort study—If applicable, explain how loss to follow-up was addressed | NA |
|  |  | Case-control study—If applicable, explain how matching of cases and controls was addressed |  |
|  |  | Cross-sectional study—If applicable, describe analytical methods taking account of sampling strategy |  |
|  |  | (e) Describe any sensitivity analyses | NA |
| Results |  |  |  |
| Participants | 13* | (a) Report numbers of individuals at each stage of study—eg numbers potentially eligible, examined for eligibility, confirmed eligible, included in the study, completing follow-up, and analysed | NA |
|  |  | (b) Give reasons for non-participation at each stage | NA |
|  |  | (c) Consider use of a flow diagram | NA |
| Descriptive data | 14* | (a) Give characteristics of study participants (eg demographic, clinical, social) and information on exposures and potential confounders | NA |
|  |  | (b) Indicate number of participants with missing data for each variable of interest | NA |
|  |  | (c) *Cohort study*—Summarise follow-up time (eg, average and total amount) | NA |
| Outcome data | 15* | *Cohort study*—Report numbers of outcome events or summary measures over time | NA |
|  |  | *Case-control study*—Report numbers in each exposure category, or summary measures of exposure | NA |
|  |  | *Cross-sectional study*—Report numbers of outcome events or summary measures | NA |
| Main results | 16 | (a) Give unadjusted estimates and, if applicable, confounder-adjusted estimates and their precision (eg, 95% confidence interval). Make clear which confounders were adjusted for and why they were included | 4 |
|  |  | (b) Report category boundaries when continuous variables were categorized | NA |
|  |  | (c) If relevant, consider translating estimates of relative risk into absolute risk for a meaningful time period | NA |
| Other analyses | 17 | Report other analyses done—eg analyses of subgroups and interactions, and sensitivity analyses | NA |
| Discussion |  |  |  |
| Key results | 18 | Summarise key results with reference to study objectives | 5-6 |
| Limitations | 19 | Discuss limitations of the study, taking into account sources of potential bias or imprecision. Discuss both direction and magnitude of any potential bias | 6 |
| Interpretation | 20 | Give a cautious overall interpretation of results considering objectives, limitations, multiplicity of analyses, results from similar studies, and other relevant evidence | 7 |
| Generalisability | 21 | Discuss the generalisability (external validity) of the study results | 7 |
| Other information |  |  |  |
| Funding | 22 | Give the source of funding and the role of the funders for the present study and, if applicable, for the original study on which the present article is based | NA |

\*Give information separately for cases and controls in case-control studies and, if applicable, for exposed and unexposed groups in cohort and cross-sectional studies.
**Note:** An Explanation and Elaboration article discusses each checklist item and gives methodological background and published examples of transparent reporting. The STROBE checklist is best used in conjunction with this article (freely available on the Web sites of PLoS Medicine at http://www.plosmedicine.org/, Annals of Internal Medicine at http://www.annals.org/, and Epidemiology at http://www.epidem.com/). Information on the STROBE Initiative is available at www.strobe-statement.org.

Data acquisition is accomplished through Python, a programming language. Data calculations are conducted using Haskell, a functional programming language.

## Results

Data from DrugBank API were accessed on June 6^th^, 2020. DrugBank did not yield a result for mefenamic acid and rimegepant. As such, these medications are removed from our study. The result is a list of 44 medications. (Table 1) We derived 946, 13244, and 135751 unique combinations for L_2_, L_3_, and L_4_, respectively. This is in accordance with the mathematical formula _44_C_*n*_, where *n* is 2, 3, and 4.

A total of 192 combinations of abortive medications are without any interaction. These are presented in Appendix 1. However, since any four-drug combination contains, as subsets, multiple two-drug and three-drug combinations, and any three-drug combinations contains, as subsets, multiple two-drug combinations, this list can be simplified to Table 2. In other words, each member of Table 2 contains medications that can be taken 2, 3, or 4 without interaction. (For example, consider the first item of Table 2: one can use any combinations of acetaminophen, lasmiditan, methylergonovine, and prednisone without any known interaction.)

Of the list of 192, 123 (64.0%) are two-drug combinations, 67 (34.9%) are three-drug combinations, and 2 (1.0%) are four-drug combinations. Therefore, 13.0% of the all possible two-drug combinations, 0.5% of all three-drug combinations, and 0.001% of the all four-drug combinations are without interactions. Of the three-drug combinations, 59 combinations include ubrogepant. The remaining 8 non-ubrogepant containing three-drug combinations include acetaminophen, prednisone, or both.

The most commonly included drugs are ubrogepant (96 inclusions), prochlorperazine (41 inclusions), tizanidine (37 inclusions), and prednisone (31 inclusions). The most commonly included medication classes are NSAIDs (133) and gepants (96 inclusions). Triptans collectively are included in only 18 combinations and 5-HT_1F_ agonists, like lasmiditan, are in 28 combinations. Haloperidol, ergotamine, methylprednisolone, and dexamethasone are in none of the non-interacting combinations.

## Discussion

This is the first known instance of an exhaustive drug interaction study. Here, we present a complete list of non-opioid, non-barbiturate containing headache abortive medication combinations without any drug-drug interactions. Besides being an exploratory and proof of concept study, our intention is also to provide headache practitioners with a list of safe and rational combination therapy.

Headache providers often use acute medications off-label. As our interest is precisely in such polypharmacy in the real-world setting, we have decided to use Jefferson Headache Manual as our source of input; the manual includes a broad list of acute medications, including commonly used neuroleptics as well as muscle relaxers, such as tizanidine. As expected, this list of oral acute medications represents a superset of level A and level B evidenced medications recommended by the American Headache Society guideline with the exception of opioids and isometheptene containing analgesics.^1^

Our study confirms the hypothesis that polypharmacy requires judicious choice of medication pairing. The majority, 87% to be exact, of randomly selected two-drug combinations produce some sort of interaction. Furthermore, our study shows that randomly selecting three-drug combinations has a 99.5% chance of producing an interaction.

Specific classes of medication also present in specific patterns when multi-drug interactions are taken into account. We will detail selected medication classes in the following sections.

### NSAIDs

NSAIDs represent a commonly used class of medication for headache treatment. Our study supports its use in combination with other abortive medications. Indeed, the majority of three-drug combinations contain one NSAID. A number of NSAIDs are safe when combined with ubrogepant or with neuroleptics. It is worth noting, however, that of the only two viable four-drug combinations, no NSAID is included.

### Ubrogepant/Gepants

Ubrogepant is the most commonly occurring medication in our list. Therefore, it is the safest medication when used in combinations. Ubrogepant combinations also comprise of the majority of viable three-drug combinations. Therefore, it can be argued that the addition of gepants to the migraine armamentarium allows for the possibility of a large number of three-drug combinations. Although rimegepant has been excluded from our search given lack of data in DrugBank, we suspect that that CGRP small molecule antagonists as a class may emerge as safe abortive options rivaling NSAIDs in the future.

### Triptans

Triptans rank low in Table 1, signifying that they are likely to interact with other medications when used in combinations. Therefore, triptans, surprisingly, are among the medication class with the most likely interaction when used in combination therapy. The safest triptan when used in combination is naratriptan, occurring in 5 combinations.

### Neuroleptics

Neuroleptics ranking varies widely. Prochlorperazine, for example, is the second safest when used in combination (Table 1). Olanzapine appears also to be well tolerated when used in combination. Haloperidol, however, inevitably produces interactions when used in combination.

## Limitations

Our study is limited by our choice of inclusion medications. Although the Jefferson Headache Manual encompasses a wide range of medications commonly used in headache medicine, it does not contain all possible medications. We believe that any of our headache colleagues will be able to show us a favorite medication that is not included in our study. However, compiling such a list is beyond the scope of our project. Indeed, our goal is an exploration of a methodological approach: we hope to show both the utility of a combinatorial approach when applied to medication interactions and to presenting a pragmatic list of useful medication combination for clinicians. Our study is also limited by our choice of excluded medications. At the outset, we are interested only in non-opioid and non-barbiturate containing analgesics as a first assumption. While inclusion of opioid and barbiturates may broaden our project, we are unsure whether it would add any value for the result of our study; opioids and barbiturates have not been endorsed by the headache community to begin with and certainly not endorsed to be used in combination.

Our method of interrogating DrugBank does not account for severity of drug interactions. For instance, acetaminophen and ubrogepant do indeed interact, leading to potential elevation of the latter drug. However, this is in stark contrast to concurrent use of prednisone with NSAIDs, which potentiate the risk for gastrointestinal bleeding.^6^ In other words, having a drug interaction does not imply that the interactions are themselves serious or warrant discontinuation. There is a wide range of adverse reactions, some are more tolerable than others and severity may vary from person to person.

Likewise, this study did not evaluate whether or not the duration of drug combination use has an effect on the risk of an adverse outcome. Most acute therapies are used in a limited capacity and we hypothesize that if a drug combination is used more frequently, then there is a higher risk of serious interaction.

## Future directions

To better understand the possible severity of drug-drug interactions, a study compiling a list of side-effects, along with the specific drug combinations, could be useful. Previous studies have investigated the effect of headache drugs on the cytochrome P450 pathways (CYP) as a culprit for many of these interactions.^7^ If a four-drug combination is needed, further studies may also help to shed light on which combinations provide the most tolerable side effect risk. We can also address the effects of alcohol and other non-pharmacological interactions that may confound some of the adverse reactions experienced clinically.

The list of drugs we use in this study does not include every abortive medication and future direction can be geared toward a specialized database that compares these abortives with other non-headache medications that the patient may be taking.

Finally, acute medications in headaches is only a part of the whole management pictures. To investigate the interactions between acute and prevention medication, a similar study by the Rutgers group is currently being conducted.

## Conclusions

The time spent by practitioners to review and reconcile patient’s medications can be cumbersome. A system to carefully address interactions between prescribed medicines can greatly reduce the medication reconciliation time and improve headache treatment. By providing an easily accessible list of abortive medications and their potential interactions, clinicians can more readily and effectively manage their refractory headache patients.

## Data Availability

Input for data is available to the public via DrugBank API.

## Abbreviations

API: Application Programming Interface
NSAIDs: Nonsteroidal Anti-inflammatory Drugs
FDA: Food and Drugs Administration
5HT1F: Serotonin receptor 1 F
CGRP: calcitonin gene-related peptide
CYP: cytochromes P450.

## Acknowledgements

None

## Statement of authorship

Category 1

a. Conception and Design: Pengfei Zhang
b. Acquisition of Data: Pengfei Zhang
c. Analysis and Interpretation of Data: Pengfei Zhang, Victor Kaytser

Category 2

a. Drafting and Manuscript: Pengfei Zhang, Victor Kaytser
b. Revising It for Intellectual Content: Pengfei Zhang, Victor Kaytser

Category 3

(α) Final Approval of the Completed Manuscript: Pengfei Zhang, Victor Kaytser

## Appendix 1

List of abortive medications without interactions.

1. acetaminophen,lasmiditan,methylergonovine,prednisone,
2. ubrogepant,olanzapine,methylergonovine,prednisone,
3. sulindac,ubrogepant,tizanidine,
4. sulindac,ubrogepant,olanzapine,
5. sulindac,ubrogepant,prochlorperazine,
6. tolmetin,ubrogepant,tizanidine,
7. tolmetin,ubrogepant,olanzapine,
8. tolmetin,ubrogepant,prochlorperazine,
9. aspirin,acetaminophen,lasmiditan,
10. aspirin,ubrogepant,tizanidine,
11. aspirin,ubrogepant,prochlorperazine,
12. zolmitriptan,ubrogepant,prednisone,
13. celecoxib,ubrogepant,tizanidine,
14. naratriptan,acetaminophen,prednisone,
15. naratriptan,ubrogepant,prednisone,
16. acetaminophen,lasmiditan,ibuprofen,
17. acetaminophen,lasmiditan,methylergonovine,
18. acetaminophen,lasmiditan,prednisone,
19. acetaminophen,droperidol,prednisone,
20. acetaminophen,methylergonovine,prednisone,
21. lasmiditan,methylergonovine,prednisone,
22. ubrogepant,frovatriptan,prednisone,
23. ubrogepant,eletriptan,prednisone,
24. ubrogepant,etodolac,tizanidine,
25. ubrogepant,etodolac,prochlorperazine,
26. ubrogepant,nabumetone,prochlorperazine,
27. ubrogepant,oxaprozin,tizanidine,
28. ubrogepant,oxaprozin,olanzapine,
29. ubrogepant,oxaprozin,prochlorperazine,
30. ubrogepant,diclofenac,prochlorperazine,
31. ubrogepant,diflunisal,tizanidine,
32. ubrogepant,diflunisal,olanzapine,
33. ubrogepant,diflunisal,prochlorperazine,
34. ubrogepant,dihydroergotamine,prednisone,
35. ubrogepant,ketorolac,tizanidine,
36. ubrogepant,ketorolac,prochlorperazine,
37. ubrogepant,salsalate,tizanidine,
38. ubrogepant,salsalate,olanzapine,
39. ubrogepant,salsalate,prochlorperazine,
40. ubrogepant,droperidol,prednisone,
41. ubrogepant,sumatriptan,naproxen,
42. ubrogepant,meloxicam,tizanidine,
43. ubrogepant,meloxicam,prochlorperazine,
44. ubrogepant,fenoprofen,tizanidine,
45. ubrogepant,fenoprofen,olanzapine,
46. ubrogepant,fenoprofen,prochlorperazine,
47. ubrogepant,flurbiprofen,tizanidine,
48. ubrogepant,flurbiprofen,prochlorperazine,
49. ubrogepant,ibuprofen,tizanidine,
50. ubrogepant,ibuprofen,prochlorperazine,
51. ubrogepant,tizanidine,indomethacin,
52. ubrogepant,tizanidine,meclofenamate,
53. ubrogepant,tizanidine,ketoprofen,
54. ubrogepant,tizanidine,piroxicam,
55. ubrogepant,tizanidine,prednisone,
56. ubrogepant,indomethacin,prochlorperazine,
57. ubrogepant,meclofenamate,olanzapine,
58. ubrogepant,meclofenamate,prochlorperazine,
59. ubrogepant,olanzapine,ketoprofen,
60. ubrogepant,olanzapine,methylergonovine,
61. ubrogepant,olanzapine,prednisone,
62. ubrogepant,ketoprofen,prochlorperazine,
63. ubrogepant,methylergonovine,metoclopramide,
64. ubrogepant,methylergonovine,prednisone,
65. ubrogepant,methylergonovine,promethazine,
66. ubrogepant,naproxen,prochlorperazine,
67. ubrogepant,piroxicam,prochlorperazine,
68. ubrogepant,prednisone,prochlorperazine,
69. olanzapine,methylergonovine,prednisone,
70. sulindac,lasmiditan,
71. sulindac,ubrogepant,
72. sulindac,tizanidine,
73. sulindac,olanzapine,
74. sulindac,prochlorperazine,
75. tolmetin,lasmiditan,
76. tolmetin,ubrogepant,
77. tolmetin,tizanidine,
78. tolmetin,olanzapine,
79. tolmetin,prochlorperazine,
80. aspirin,acetaminophen,
81. aspirin,lasmiditan,
82. aspirin,ubrogepant,
83. aspirin,tizanidine,
84. aspirin,prochlorperazine,
85. zolmitriptan,ubrogepant,
86. zolmitriptan,prednisone,
87. celecoxib,ubrogepant,
88. celecoxib,tizanidine,
89. naratriptan,acetaminophen,
90. naratriptan,ubrogepant,
91. naratriptan,prednisone,
92. acetaminophen,lasmiditan,
93. acetaminophen,droperidol,
94. acetaminophen,ibuprofen,
95. acetaminophen,methylergonovine,
96. acetaminophen,prednisone,
97. lasmiditan,etodolac,
98. lasmiditan,nabumetone,
99. lasmiditan,oxaprozin,
100. lasmiditan,diclofenac,
101. lasmiditan,diflunisal,
102. lasmiditan,ketorolac,
103. lasmiditan,salsalate,
104. lasmiditan,meloxicam,
105. lasmiditan,fenoprofen,
106. lasmiditan,flurbiprofen,
107. lasmiditan,ibuprofen,
108. lasmiditan,indomethacin,
109. lasmiditan,meclofenamate,
110. lasmiditan,ketoprofen,
111. lasmiditan,methylergonovine,
112. lasmiditan,naproxen,
113. lasmiditan,piroxicam,
114. lasmiditan,prednisone,
115. ubrogepant,frovatriptan,
116. ubrogepant,eletriptan,
117. ubrogepant,chlorpromazine,
118. ubrogepant,etodolac,
119. ubrogepant,almotriptan,
120. ubrogepant,nabumetone,
121. ubrogepant,oxaprozin,
122. ubrogepant,diclofenac,
123. ubrogepant,diflunisal,
124. ubrogepant,dihydroergotamine,
125. ubrogepant,ketorolac,
126. ubrogepant,salsalate,
127. ubrogepant,droperidol,
128. ubrogepant,sumatriptan,
129. ubrogepant,meloxicam,
130. ubrogepant,fenoprofen,
131. ubrogepant,flurbiprofen,
132. ubrogepant,ibuprofen,
133. ubrogepant,tizanidine,
134. ubrogepant,indomethacin,
135. ubrogepant,meclofenamate,
136. ubrogepant,olanzapine,
137. ubrogepant,ketoprofen,
138. ubrogepant,methylergonovine,
139. ubrogepant,metoclopramide,
140. ubrogepant,naproxen,
141. ubrogepant,piroxicam,
142. ubrogepant,prednisone,
143. ubrogepant,prochlorperazine,
144. ubrogepant,promethazine,
145. ubrogepant,rizatriptan,
146. frovatriptan,prednisone,
147. eletriptan,prednisone,
148. etodolac,tizanidine,
149. etodolac,prochlorperazine,
150. nabumetone,prochlorperazine,
151. oxaprozin,tizanidine,
152. oxaprozin,olanzapine,
153. oxaprozin,prochlorperazine,
154. diclofenac,prochlorperazine,
155. diflunisal,tizanidine,
156. diflunisal,olanzapine,
157. diflunisal,prochlorperazine,
158. dihydroergotamine,prednisone,
159. ketorolac,tizanidine,
160. ketorolac,prochlorperazine,
161. salsalate,tizanidine,
162. salsalate,olanzapine,
163. salsalate,prochlorperazine,
164. droperidol,prednisone,
165. sumatriptan,naproxen,
166. meloxicam,tizanidine,
167. meloxicam,prochlorperazine,
168. fenoprofen,tizanidine,
169. fenoprofen,olanzapine,
170. fenoprofen,prochlorperazine,
171. flurbiprofen,tizanidine,
172. flurbiprofen,prochlorperazine,
173. ibuprofen,tizanidine,
174. ibuprofen,prochlorperazine,
175. tizanidine,indomethacin,
176. tizanidine,meclofenamate,
177. tizanidine,ketoprofen,
178. tizanidine,piroxicam,
179. tizanidine,prednisone,
180. indomethacin,prochlorperazine,
181. meclofenamate,olanzapine,
182. meclofenamate,prochlorperazine,
183. olanzapine,ketoprofen,
184. olanzapine,methylergonovine,
185. olanzapine,prednisone,
186. ketoprofen,prochlorperazine,
187. methylergonovine,metoclopramide,
188. methylergonovine,prednisone,
189. methylergonovine,promethazine,
190. naproxen,prochlorperazine,
191. piroxicam,prochlorperazine,
192. prednisone,prochlorperazine,

